# Childhood immunological imprinting of cross-subtype antibodies targeting the hemagglutinin head domain of influenza viruses

**DOI:** 10.1101/2025.09.24.25335646

**Authors:** Shuk Hang Li, Bo Wang, Gyunghee Jo, Artem Mikelov, Reilly K. Atkinson, Valerie Le Sage, Colleen Furey, Jordan T. Ort, Naiqing Ye, Katharina Röltgen, Shilpa A. Joshi, Ji-Yeun Lee, Taylor A. Pursell, Elizabeth M. Drapeau, Julianna Han, Amy P. Callear, Ronald G. Collman, Arnold S. Monto, Emily T. Martin, Seema S. Lakdawala, Andrew B. Ward, Ian A. Wilson, Scott D. Boyd, Scott E. Hensley

## Abstract

Influenza virus cross-subtype antibodies targeting the hemagglutinin (HA) head are rare. Here, we found that a large proportion of monoclonal antibodies (mAbs) isolated from individuals immunized with the 2021-22 seasonal influenza vaccine bound to an epitope on the HA head of both the H1N1 vaccine strain and H3N2 strains from the mid-1990s. These H1/H3 cross-reactive antibodies were also found in polyclonal sera, but only in samples from individuals born in the 1990s. Ferrets sequentially exposed to an H3N2 virus from the 1990s and a contemporary seasonal influenza vaccine produced the same type of H1/H3 cross-reactive antibodies. We found evidence that H1N1 viruses are currently evolving within the human population to abrogate the binding of these antibodies. Together, our study demonstrates how prior influenza virus exposures can influence the specificity of antibodies elicited by entirely different influenza virus subtypes, and how viruses evolve to escape these types of antibodies.

## Background

Influenza viruses circulate in humans and cause annual seasonal epidemics in the Northern and Southern Hemispheres. Hemagglutinin (HA) is the most abundant protein on the surface of influenza virions and is responsible for cell attachment and entry^1,2^. Most neutralizing antibodies induced by influenza vaccinations and infections bind to the more immunodominant globular head of HA and sterically block virus attachment to host cells^3^. However, the HA head of human influenza viruses continually acquires amino acid substitutions and glycan changes that abrogate antibody binding through a process called ‘antigenic drift’^4^. This process leads to seasonal epidemics and necessitates the routine update of influenza vaccine formulations that are antigenically matched to circulating viruses.

Influenza A subtypes can be divided into two phylogenetic groups, group 1 and group 2, based on the HA sequence and antigenic properties^5–8^. The HA head differs greatly among subtypes both within and across antigenic groups, despite housing the receptor binding site.

Conversely, epitopes within the lower part of HA, called the HA stalk, are mostly conserved among subtypes within an antigenic group^9^. Rare monoclonal antibodies (mAbs) bind to the HA stalk of both group 1 and group 2 viruses^10–22^. Stalk-targeting mAbs can block the viral fusion machinery of HA and also provide protection via non-neutralizing Fc gamma receptor-dependent mechanisms *in vivo*^10,23^. Cross-reactive antibodies targeting the HA head of different influenza virus subtypes are even rarer; however, some mAbs targeting HA head epitopes in group 1 and 2 viruses have been recently identified^24–32^.

Early childhood influenza virus exposures prime memory B cells that can be recalled by antigenically distinct viral strains later in life^33^. This phenomenon, initially called “original antigenic sin” in 1960^34^, has been more commonly referred to as ‘immune imprinting’ in the 21^st^ century^35–37^. H1N1 (group 1) and H3N2 (group 2) viruses have co-circulated in humans since 1977. Therefore, it is important to determine how prior exposure to one viral subtype affects the development of immune responses against subsequent exposure to the second viral subtype.

Studies have extensively evaluated how prior H1N1 exposures impact immunity to antigenically distinct H1N1 strains^12,38–42^, but few studies have examined how immune imprints established by H3N2 infections affect immune responses elicited by subsequent H1N1 exposures^43–45^. We previously characterized antibody responses in ferrets sequentially exposed to H3N2 and H1N1 viruses^46^. We found that HA stalk antibodies were elicited by initial infections and then boosted upon subsequent infections with heterosubtypic viral subtypes. We observed minor cross-subtype boosting of polyclonal antibodies reactive to the HA head in these studies, but we did not investigate the specificity or functionality of these antibodies.

Here, we produced a large panel of mAbs from plasmablasts isolated from 17 individuals who received the 2021-22 seasonal influenza vaccine. Unexpectedly, we found that many H1N1 vaccine-reactive mAbs bound strongly to the HA head of H3N2 viruses that circulated in the 1990s. Most of these cross-subtypic mAbs were isolated from individuals born in the 1990s, which led us to hypothesize that H3N2 infections ∼30 years ago primed B cell responses that could later be recalled by contemporary H1N1 viruses. To address this hypothesis, we completed comprehensive serological analyses with serum from individuals with different birth years, and we also tested influenza vaccines in ferrets previously exposed to distinct H3N2 strains. Finally, we characterized the fine-specificity and functionality of these cross-subtype antibodies targeting the globular HA head of historical H3N2 and contemporary H1N1 viruses.

## Results

### Many H1-reactive mAbs elicited by the 2021-22 vaccine bind to H3 from the mid-1990s

We collected peripheral blood mononuclear cells (PBMCs) from 17 individuals seven days after receiving the 2021-22 FluLaval inactivated egg-based influenza vaccine. We single-cell sorted plasmablasts from the PBMCs, sequenced their heavy and light chain antibody genes, and recombinantly expressed 297 mAbs. We established a multiplex binding assay to determine if these mAbs bound to HAs from the vaccine strains and historical viruses. In these assays, we tested mAb binding to a panel of recombinant full-length HAs representative of influenza viruses that circulated over the past century. In total, we measured mAb binding to HAs from the four components of the 2021-22 egg-based influenza vaccine, HAs from 27 viruses that circulated from 1918 to 2014 (9 H1N1 viruses, 10 H3N2 viruses, and 8 influenza B viruses (IBVs)), and ‘headless’ H1 and H3 proteins so that we could measure mAb binding to the HA stalk domain (**Figure S1A**).

We found that 65 of the mAbs bound to at least one vaccine strain HA (**Figure S1A-S1B**). Thirty-one (47.7%) of these mAbs bound to only the H1N1 vaccine HA (A/Victoria/2570/2019), 11 (16.9%) to the H3N2 vaccine HA (A/Tasmania/503/2020), 9 (13.8%) to either or both IBV vaccine strains (B/Washington/02/2019 or B/Phuket/3073/2013), and 14 (21.5%) to more than one vaccine HA (**Figure S1B)**. Consistent with our previous studies^41,42,47^, all vaccine-reactive mAbs bound to one or more HAs from historical viruses of the same subtype (**Figure S1A**). For example, every mAb that bound to the HA of the H1 vaccine strain A/Victoria/2570/2019 also bound to an HA from one or more historical H1N1 viruses.

Surprisingly, many of the mAbs that bound to the H1 vaccine strain also bound to HAs from H3N2 viruses that circulated in the 1990s (A/Stockholm/20/1993 and/or A/New York/631/1996; **Figure S1A**). Most previously described cross-subtype mAbs bind to the stalk domain of HA; however, only one of these H1/H3 cross-reactive mAbs in our study bound to headless HA proteins in our assays (**Figure S1A**). This finding suggests that most of these H1/H3 cross-reactive mAbs here target the more divergent HA head domain.

### Most H1/H3 cross-subtype mAbs neutralize contemporary H1N1 vaccine strain

We completed additional analyses of the H1 vaccine-reactive mAbs that unexpectedly bound to HAs from historical H3N2 viruses. Notably, most of the participants in our study were born in the 1990s, and most of the cross-subtype mAbs bound to the HAs from H3N2 viruses that circulated in the 1990s. We grouped the H1-reactive mAbs based on their cross-reactivity with HAs from historical H3N2 viruses: Group A mAbs have narrow back-reactivity to only an H3 HA from 1996 (A/New York/631/1996), Group B mAbs have broader back-reactivity to multiple H3 HAs, and Group C mAbs are conventional and do not bind to any of the H3 HAs that we tested (**Figure 1A**). Although most cross-subtype mAbs isolated from the same participant represented independent B cell lineages with distinct VDJ gene usage, several of the mAbs were derived from the same lineages. For instance, three Group A mAbs from participant 88 (88_B4, 88_C11, 88_D10) were derived from the same clone, whereas the six Group B mAbs from participant 33 included both unrelated lineages and clonally related members. In total, 15 of the mAbs could be assigned to expanded clones, highlighting that our dataset captures both single representatives of diverse B cell lineages and multiple members of specific clonal families.

**Figure 1.**
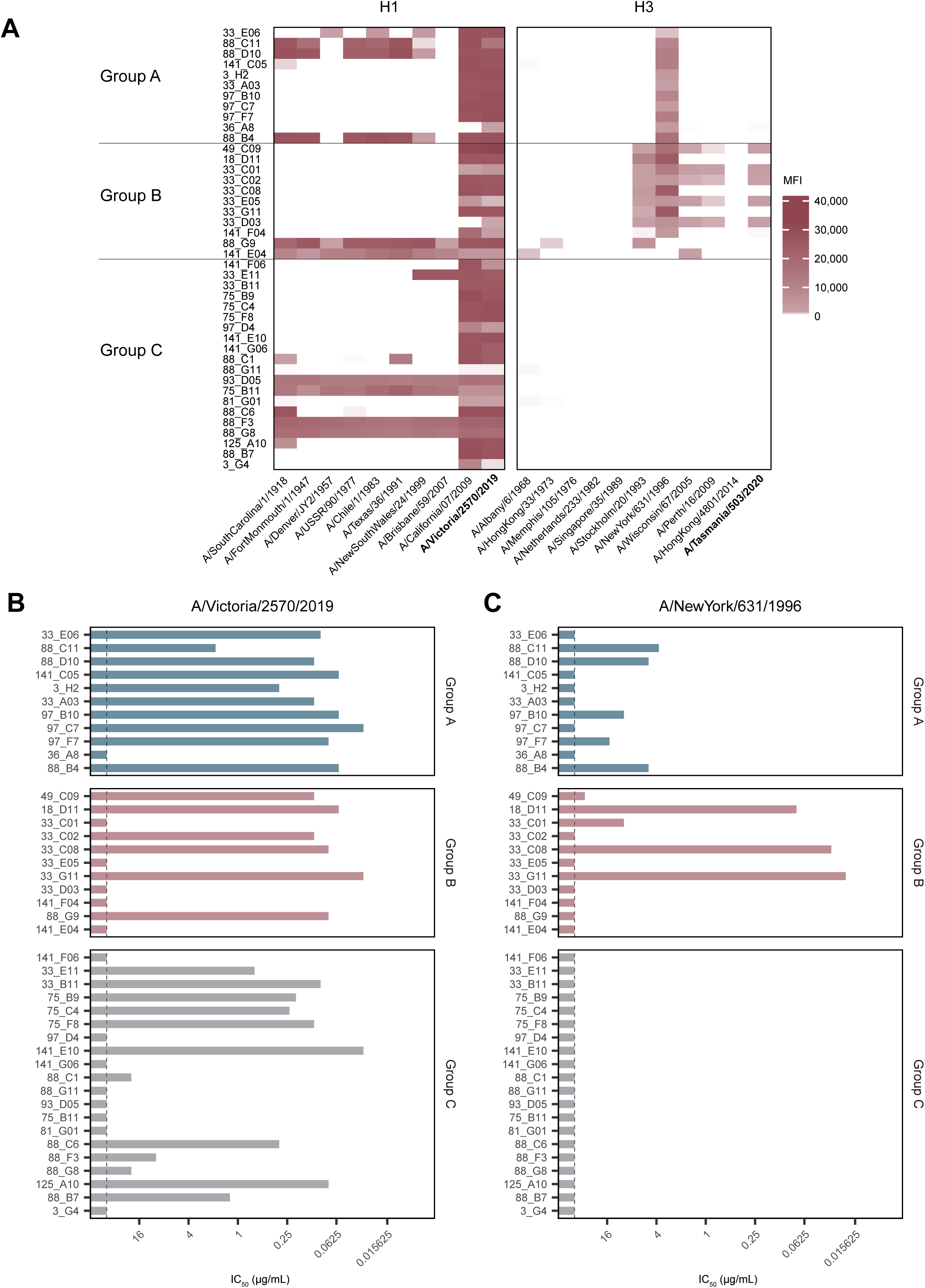
Influenza vaccine-elicited antibodies cross-react to HAs from the 2021-22 H1N1 vaccine strain and H3N2 strains from the mid-1990s. Monoclonal antibodies isolated seven days after 2021-22 inactivated influenza vaccination were measured against a panel of H1 and H3 HAs, including representative historical strains and the vaccine strains (H1N1 A/Victoria/2570/2019 and H3N2 A/Tasmania/503/2020), in a multiplexed binding assay (**A**). The intensity of binding is shown in mean fluorescence intensity (MFI). H1N1 vaccine-reactive antibodies are categorized into three groups: Group A antibodies have narrow reactivity to H3N2 HA (n = 11); Group B antibodies have broader back reactivity to H3N2 recognizing more than one H3 HA (n = 11); and Group C antibodies (n = 20) do not have any specificity against H3 HAs. We measured neutralization of these H1 vaccine-reactive antibodies against the H1N1 vaccine strain A/Victoria/2570/2019 (**B**) or the 1996 H3N2 strain A/New York/631/1996 (C). 50% inhibitory concentration values were plotted (IC_50_). Data represent the mean of two independent experiments. Dashed lines indicate the limit of detection in (**B** and **C**).

Ten of the 11 Group A mAbs neutralized the H1 vaccine strain A/Victoria/2570/2019, whereas just over 50% of Group B and Group C mAbs neutralized the H1 vaccine strain (**Figure 1B**). Several Group A and B mAbs, but not Group C mAbs, neutralized an H3N2 virus from 1996 (A/New York/631/1996; **Figure 1C**). Group A mAbs did not neutralize A/New York/631/1996 H3N2 viruses as efficiently as the H1N1 vaccine strain, while some of the Group B mAbs had similar neutralizing potency against both the H1 vaccine strain and the 1996 H3N2 strain (compare **Figures 1B** and **1C**). These data indicate that many mAbs isolated from individuals after influenza vaccination target neutralizing epitopes that seem to be conserved on the globular HA heads of contemporary H1N1 vaccine strains and historical H3N2 viruses.

### H1/H3 cross-reactive mAbs target an HA head epitope involving residue 145

We performed experiments to better understand the binding epitopes of Group A mAbs that narrowly react to an H3N2 from 1996 and Group B mAbs that have broader reactivity to historical H3N2 viruses. Koel and colleagues demonstrated that an N145K HA substitution that arose in H3N2 viruses in the mid-1990s led to a major antigenic change^48,49^. Most H3N2 viruses that circulated between ∼1990 and 2004, including the A/New York/631/1996 H3N2 virus in our study, possessed a lysine (K) at HA residue 145 (**Figure S2**, dark blue). Coincidentally, most pandemic H1N1 (pH1N1) viruses, including the 2021-22 H1 vaccine strain, also possess a lysine at position 145 (H3 numbering).

We hypothesized that the H1/H3 cross-reactive mAbs in our study target an HA head epitope involving a lysine at residue 145, which is conserved between mid-1990s H3s and contemporary H1s. To address this hypothesis, we introduced a K145N substitution into the HA of the A/New York/631/1996 H3N2 and tested mAb binding to both the wildtype and mutant HAs. Group A mAbs bound to the native 1996 H3 HA with K145 but were unable to bind to the modified 1996 H3 with N145 (**Figure 2A**). The K145N substitution also reduced binding of all Group B mAbs, but some of these mAbs maintained weak binding to the mutant HA. As expected, Group C mAbs bound to neither H3 HA. We then completed a complementary experiment by measuring mAb binding to the HA of A/Stockholm/20/1993 H3N2 engineered to possess an N145K substitution. Consistent with our initial experiments (**Figure 1A**), Group A mAbs didn’t bind to the native 1993 HA with an N145; however, these mAbs bound to the 1993 HA engineered with a lysine at residue 145 (**Figure 2B**). The N145K substitution also increased binding of most Group B mAbs to 1993 H3 HA (**Figure 2B).**

**Figure 2.**
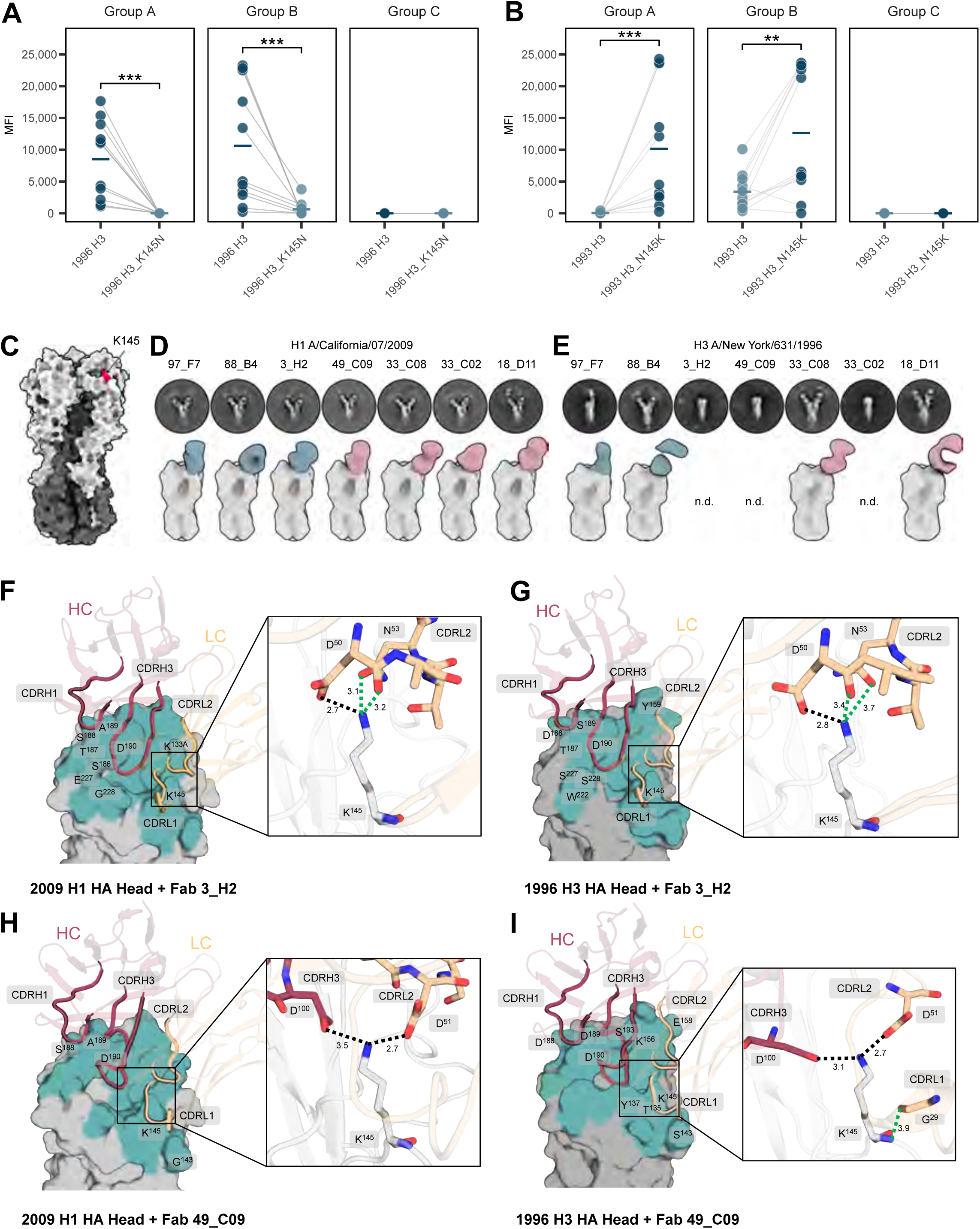
H1/H3 cross-subtype mAbs target an epitope involving residue 145. 1996 H3 HA (A/New York/631/1996; WT K145) or 1993 H3 HA (A/Stockholm/20/1993; WT N145) with either asparagine (N) or lysine (K) at amino acid 145 were generated using site-directed mutagenesis. Binding of H1 vaccine-reactive antibodies to either the WT or the mutant HAs in the 1996 H3 backbone (**A**) or in the 1993 H3 backbone (**B**) was measured in a multiplexed binding assay. MFIs represent the mean of two independent experiments. Horizontal bars indicate means, and statistical analyses were done using a Wilcoxon signed-rank test (**A** and **B**). **P < 0.01, ***P < 0.001. (**C**) Structure of A/California/04/2009 H1 HA (PDB: 4M4Y) with residue K145 highlighted in red on the head domain. Negative-stain EM analysis of seven Group A and B mAbs bound to A/California/07/2009 H1 HA (**D**) or A/New York/631/1996 H3 HA (**E**). Representative 2D class averages (top) and composite maps showing Fab density aligned to HA (bottom) are shown. For mAbs that did not bind to H3 (n.d. indicates structures not determined), only 2D class averages of unbound HA are shown. Crystal structures of Fab-HA complexes: A/California/04/2009 H1 HA with Fab 3_H2 (**F**), A/New York/631/1996 H3 HA head with Fab 3_H2 (**G**), A/California/04/2009 H1 HA with Fab 49_C09 (**H**), A/New York/631/1996 H3 HA head with Fab 49_C09 (**I**). The HA head domains are shown in surface representation in grey, with the epitope regions (defined as residues with a buried surface area > 0 Å^2^) highlighted in turquoise. Antibodies are shown in cartoon representation, with heavy chains colored in dark red and light chains colored in wheat. Labeled residues indicate those involved in hydrogen bonds or salt bridge formation upon HA engagement. Insets on the right display zoomed-in views of the interactions involving K145 for each complex. Green dashed lines represent hydrogen bonds, black dashed lines represent salt bridges, which also involve hydrogen bonding in the four crystal structures presented here.

We used negative-stain electron microscopy (nsEM) to visualize the binding interactions of seven Group A and B mAbs (Group A in blue: 97_F7, 88_B4, and 3_H2; Group B in pink: 49_C09, 33_C08, 33_C02, and 18_D11) to the HA of A/California/07/2009 H1N1 and A/New York/631/1996 H3N2 viruses (**Figure 2C-E**). All seven mAbs bound to an epitope in the HA head of A/California/07/2009 H1N1 directly involving or adjacent to residue 145 (**Figure 2C-D**). For A/New York/631/1996 H3N2 HA, Fab binding was detected in 2D classifications for four of the seven mAbs (97_F7 and 88_B4 from Group A, and 33_C08 and 18_D11 from Group B) (**Figure 2E**), consistent with their relatively higher H3N2 neutralizing potency compared to some other Group A and B mAbs (**Figure 1C**). These four antibodies showed similar binding orientations with very minor differences when bound to H1 and H3 HAs.

To further investigate the structural basis underlying the binding patterns of Group A and B mAbs, we determined crystal structures of Fabs of a Group A mAb (3_H2) and a Group B mAb (49_C09) complexed to HAs from H1N1 (A/California/04/2009) and H3N2 (A/New York/631/1996) viruses (**Figure 2F-I**). In addition, a crystal structure of 49_C09 was determined in complex with the H1 vaccine HA (A/Victoria/2570/2019) (**Figure S3A**). Consistent with our site-directed mutagenesis and nsEM analyses, we found that the binding footprint of the Group A and B mAbs included the lysine at HA residue 145. Aspartic acid residues on light chain complementarity-determining region 2 (CDRL2) of both mAbs and heavy chain complementarity-determining region 3 (CDRH3) of 49_C09 were critical for the interaction with K145 (**Figure 2F-I**). The CDRL1 of 49_C09 also contributed a hydrogen bond to the recognition of 1996 H3 HA via K145 (**Figure 2I**). Additionally, both mAbs primarily recognized the 180-loop of HA at the end of the 190-helix via CDRH1 and CDRH3 (**Figure 2F-I**), gripping the loop in a vice-like manner. The CDRH3 of both mAbs contributed to the HA head recognition through its insertion into the receptor binding site, also facilitating interactions of the CDRL2 (both mAbs) with the 150-loop and K145 (**Figure 2F-I, Figure S3B-C**). Thus, K145 serves as a critical residue enabling both Group A and Group B mAbs to exhibit cross-subtype reactivity.

Interestingly, the Group A mAb 3_H2 engaged HA K145 primarily via its light chain (CDRL2), forming hydrogen bonds and a single salt bridge (**Figure 2F-G**). In contrast, the Group B mAb 49_C09 interacted with K145 with both heavy (CDRH3) and light (CDRL2, CDRL1) chains by forming hydrogen bonds and a pair of salt bridges (**Figure 2H-I)**. The longer CDRH3 of Group B mAb allowed it to insert more deeply into the confined space of the receptor binding site (**Figure 2H-I**), enabling more extensive interactions with surrounding residues, including K145. Notably, the increased buried surface area (BSA) for CDRH3 of the Group B Fab suggests that the antibody forms more extensive contacts when bound to H3 HA compared to H1 HAs (**Figure S3D**). In contrast, no noticeable BSA differences were observed in the Group A Fab complex structures for H1 and H3 HAs. Thus, the Group B mAb seems to possess a certain degree of adaptability in binding to broader HA subtypes.

Although K145 is an important residue for mAb recognition of the HA head, other HA residues also play an essential role in the cross-reactivity to both H1 and H3. In addition to K145, the epitope residues (defined as HA residues with BSA > 0 Å^2^) W153, K156, S186, T187, D190, S193, and L194 are conserved between the 2009 H1 and 1996 H3 HAs, and some of these residues also form hydrogen bonds or salt bridges with both mAbs (**Figure S3B-C**).

Residues that differed between the H1 and H3 head also helped stabilize the interaction with these mAbs. For example, S188 in 2009 H1 HA, I188 in 2019 H1 HA, and D188 in 1996 H3 HA interacted with both mAbs through hydrogen bonds (**Figure S3B-C**). The K133A insertion in 2009 H1 HA, but not in 1996 H3 HA, made hydrogen interactions only with the Group A mAb (**Figure S3B**). Furthermore, W222 of 1996 H3 HA provides an extra hydrophobic surface to interact with the Group A mAb (**Figure S3B**).

The additional interactions of the Group B mAb may increase the antibody’s ability to bind to earlier H3 HAs without K145. For example, G143 in 2009 H1 HA and S143 in 1996 H3 HA are both capable of interacting with residue G68 on framework region 3 of the Group B mAb’s light chain through hydrogen bonds to enhance binding (**Figure S3C**). The light chain is also indispensable in recognizing K145 in both antibodies (**Figure 2F-I**). The interactions with K145 driven by the light chain are negatively affected when substituted by N145 (as in the A/Stockholm/20/1993 H3 HA) due to the loss of hydrogen bonds and salt bridges (**Figure S3B-C**) or other substitutions. However, in the Group B mAb-H3 HA structure, the increased BSA contributed by the heavy chain seems to partially compensate for K145 replacement in other H3 viruses (**Figure S3D**). Collectively, these structures show that Group A and B mAbs target an epitope adjacent to the HA receptor binding site that is partially conserved between contemporary H1N1 and 1990s H3N2 viruses and includes HA residue K145.

### Evidence that H1/H3 cross-reactive antibodies were primed by H3N2 viruses in the 1990s

We hypothesized that childhood exposures to H3N2 viruses that had an HA with K145 in the 1990s primed cross-subtype HA antibody responses that can be recalled by contemporary H1N1 viruses that also have an HA with K145. H3N2 with HA K145 circulated sporadically since the late 1970s, dominating circulation between 1990 and 2004 (**Figure S2**). We completed absorption experiments to determine if H1/H3 cross-subtype HA antibodies are more prevalent in individuals born before 2004 than those born after 2004, when H3N2 viruses with HA K145 no longer circulated. We tested serum samples collected from 80 individuals with different birth years after vaccination. Absorptions with beads coupled with an HA from a 1996 H3N2 virus (that possessed K145) removed the majority of H1-reactive serum antibody from four individuals born in the 1990s, but did not greatly affect H1-reactive antibody levels in the serum of individuals born after 2004 (**Figure 3A-B**). Absorptions with beads coupled with the same 1996 H3 HA that was engineered to possess N145 did not remove as much H1-reactive serum antibody from the same four individuals born in the 1990s (**Figures S4A**), indicating that some of these cross-subtype H1/H3 polyclonal antibodies targeted an HA epitope involving K145.

**Figure 3.**
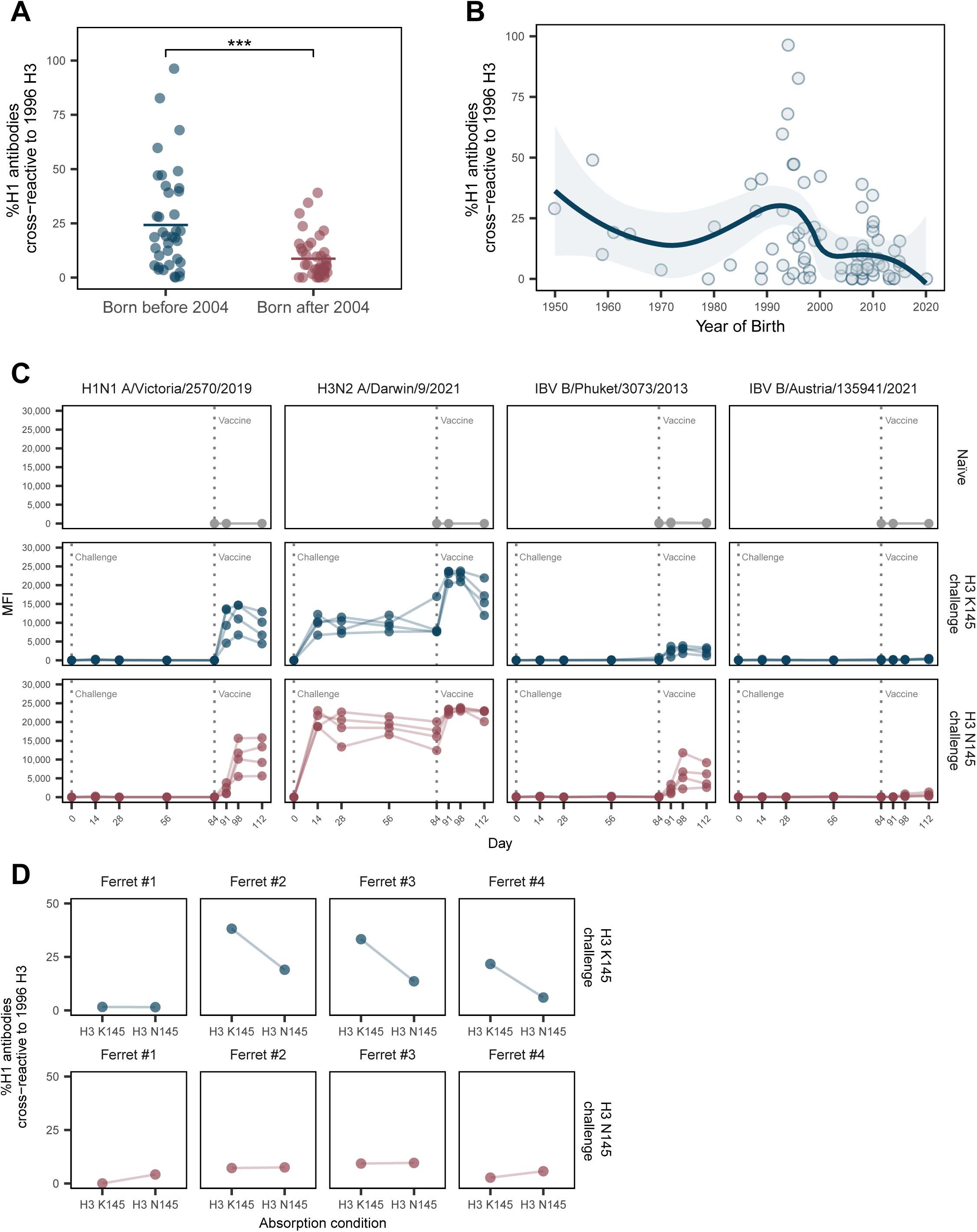
Polyclonal H1/H3 cross-reactive antibodies are enriched in individuals born in the 1990s and can be elicited in ferrets primed with H3N2 with K145. Serum binding to H1 vaccine HA (A/Victoria/2570/2019) was measured after incubating with HA-coupled beads to remove antibodies reactive to 1996 H3 HA (A/New York/631/1996) in individuals born before 2004 (blue) and individuals born after 2004 (red) (**A**). Antibody titers are represented as percentages of H1/H3 cross-reactive antibodies by measuring the reduction of H1 vaccine-reactive antibodies after removal of 1996 H3 HA-reactive antibodies. (**B**) Percent H1/H3 cross-reactive antibodies in individuals are graphed as a function of year of birth. (**C**) Ferrets (n = 4 per group) were infected with A/Nanchang/933/1995 H3N2 with HA K145 (blue) or A/Nanchang/933/1995 with HA N145 (red) and vaccinated 84 days later with a half-dose human FluLaval influenza vaccine. Naïve ferrets were not primed with any virus and only received the vaccine on day 84 (grey). Binding against the HA of the four influenza vaccine components (H1N1 A/Victoria/2570/2019, H3N2 A/Darwin/9/2021, IBV B/Phuket/3073/2013, and IBV B/Austria/135941/2021) was quantified in a multiplexed binding assay. (**D**) Absorption assays were performed to determine the amount of H1/H3 cross-reactive antibodies in ferrets primed with A/Nanchang/933/1995 H3N2 with HA K145 (blue) or A/Nanchang/933/1995 with HA N145 (red) that target either epitope involving K145 or other conserved epitopes between H3 and H1 HA. Data are representative of two independent experiments. Horizontal bars in (**A**) indicate means. The trend line in (**B**) is a locally estimated scatterplot smoothing curve (smoothing parameter = 0.6) with 95% CIs. Data in (**A**) were analyzed using a Wilcoxon rank-sum test. ***P < 0.001.

These data indicate that H1/H3 cross-reactive antibodies that recognize an epitope involving HA residue 145 can be detected in polyclonal sera and are at high levels in a subset of individuals born in the 1990s.

Next, we completed a series of ferret studies to determine if H3N2 infections can prime H1/H3 cross-subtype K145-reactive HA antibodies. We infected ferrets with either A/Nanchang/933/1995 H3N2 (that has HA K145) or A/Nanchang/933/1995 H3N2 that was engineered to have an asparagine at HA residue 145 (N145). We then vaccinated the ferrets with the FluLaval inactivated egg-based influenza vaccine 84 days after H3N2 infection. As a control, we included a group of ferrets that only received the vaccine without prior influenza virus exposure.

Ferrets infected with the H3N2 A/Nanchang/933/1995 with HA K145 or the A/Nanchang/933/1995 with HA N145 produced antibodies that bound to HA from both versions of H3 HA (**Figure S4B**, blue and red). These infection-elicited antibodies cross-reacted to the H3 vaccine strain A/Darwin/9/2021, but did not react to the H1 or IBV vaccine strains (**Figure 3C**). Ferrets pre-exposed to either H3N2 virus and subsequently immunized produced antibodies that strongly reacted to the H1 and H3 vaccine HAs, with lower reactivity to the IBV vaccine HAs (**Figure 3C**, blue and red). In contrast, the vaccine elicited weak antibody responses in ferrets without prior exposure (**Figure 3C**, grey). These data suggest that H3N2 exposure primed ferrets to produce antibodies that react to both H1N1 and H3N2 and, to a lesser extent, to IBV.

Interestingly, H3N2-exposed animals, but not previously naïve animals, produced high levels of H1N1, H3N2, and IBV-reactive antibodies following vaccination. It is unclear if this is due to H3N2 priming of cross-reactive CD4^+^ T cell responses that broadly provided help to B cells recognizing the different vaccine antigens, or if H3N2 directly primed cross-subtype reactive B cells that could be recalled by vaccine antigens^50^. To address this, we performed absorption assays to investigate whether the H1-reactive antibodies in the sera from ferrets after vaccination were cross-reactive to H3 HA. Three of the four ferrets that were previously exposed to A/Nanchang/933/1995 with HA K145 produced high levels of H1-reactive antibodies that cross-reacted to H3 HA with K145 (**Figure 3D**, blue). These polyclonal antibodies did not cross-react as well to the H3 HA with N145, suggesting that many of these antibodies were directed against an HA epitope involving lysine at residue 145. It is unclear why only three of the four ferrets previously exposed to A/Nanchang/933/1995 with HA K145 had antibodies with this phenotype. In contrast, all four of the animals that were previously exposed to A/Nanchang/933/1995 with HA N145 produced low levels of H1/H3 cross-reactive antibodies following vaccination (**Figure 3D**, red). These data suggest that B cells primed by H3N2 HAs with K145 from the 1990s can be recalled by recent H1N1 vaccine strains.

### H1N1 viruses recently acquired a substitution that abrogates the binding of H1/H3 cross-subtype antibodies

Given that many H1/H3 cross-reactive antibodies are potently neutralizing (**Figure 1B-C**) and common in a subset of adults (**Figure 3A-B**), we next asked if recent H1N1 viruses have evolved to escape these antibody responses. Mutations at HA residue 145 have been sporadically detected in pH1N1 viruses since 2009, and a large fraction of H1N1 viruses that have circulated since the 2022-23 season had an HA with a K145R substitution (**Figure 4A**). We tested binding and neutralization of the 42 H1-reactive mAbs elicited by the 2021-22 vaccine to A/Minnesota/35/2022, a recent H1N1 strain with the K145R HA substitution. Group A mAbs that have narrow back reactivity to mid-1990s H3N2 viruses (**Figure 1A**) bound to and neutralized the H1 vaccine strain but neither bound nor neutralized the drifted A/Minnesota/35/2022 H1N1 virus (**Figure 4B-C**). Group B mAbs that have broader back reactivity to multiple H3 HAs (**Figure 1A**) also bound and neutralized the drifted A/Minnesota/35/2022 H1N1 virus less efficiently compared to the H1N1 vaccine strain (**Figure 4B-C**). Some Group C mAbs bound to and neutralized the drifted H1N1 virus, while others had major reductions in binding and neutralization (**Figure 4B-C**), likely because A/Minnesota/35/2022 H1N1 virus possesses HA substitutions at residues other than 145 (**Figure S5**, residues in blue).

**Figure 4.**
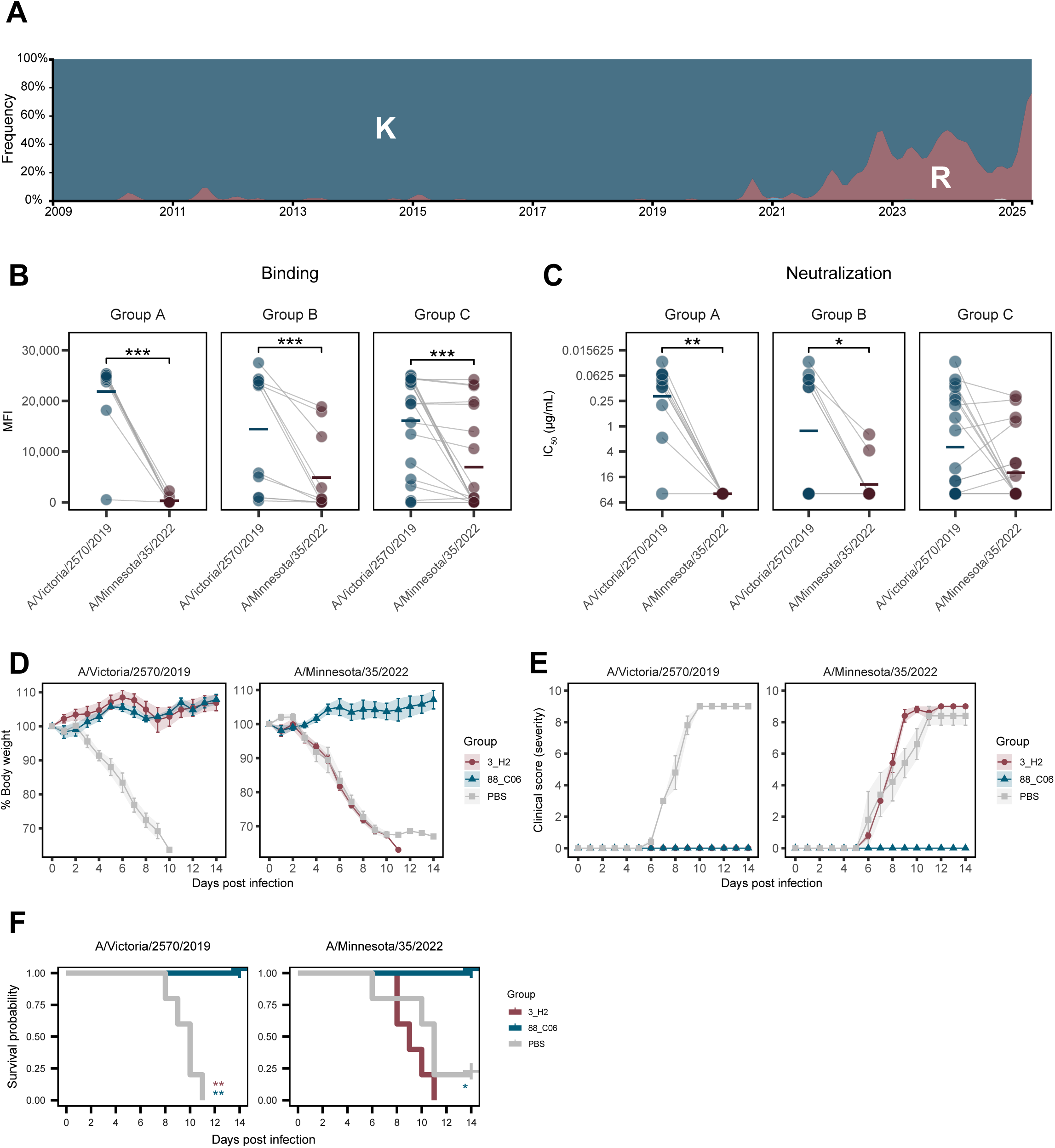
H1/H3 cross-subtype antibodies do not bind to circulating H1N1 strains that acquired a mutation at residue 145. (**A**) Frequency of amino acids at residue 145 in H1N1 HA is shown. H1N1 strains with K145R substitution started to circulate around 2022. Binding (**B**) and neutralization (**C**) of H1 vaccine-reactive mAbs were measured against the H1N1 vaccine strain (A/Victoria/2570/2019; blue) and a circulating strain with K145R mutation (A/Minnesota/35/2022; red). Group A mAb 3_H2 (red), Group C mAb 88_C06 (blue), or PBS (grey) were injected intraperitoneally into DBA/2 mice (n = 5 per group), and mice were infected intranasally with either A/Victoria/2570/2019 or A/Minnesota/35/2022 six hours later. Body weights (**D**), clinical scores that indicate disease severity (**E**), and survival (**F**) were monitored for 14 days after infection. Data represent the mean of two independent experiments and were analyzed using a Wilcoxon signed-rank test (**B** and **C**). Horizontal bars indicate means (**B**) or geometric means (**C**). Mouse experiments are representative of three independent experiments. Data are represented as means ± SEMs in (**D** and **E)**. Data in (**F**) were analyzed by a log-rank test (Mantel-Cox) comparing differences in survival to the control group receiving PBS only before infection. *P < 0.05, **P < 0.01, ***P < 0.001.

We passively transferred a Group A H1/H3 cross-subtype mAb or a Group C mAb that did not target the cross-subtype H1/H3 epitope into DBA/2 mice to determine if they could protect against the recently drifted H1N1 virus. As a control, we passively transferred PBS into animals prior to infection. Mice that received PBS prior to either A/Victoria/2570/2019 (H1N1 vaccine strain) or A/Minnesota/35/2022 (drifted H1N1) rapidly lost weight, showed clinical signs, and died or needed to be euthanized by 11-13 days after infection (**Figure 4D-4F**, grey). As expected, mice receiving the Group C mAb 88_C06 were fully protected against infection with both the vaccine strain and drifted H1N1 strain (**Figure 4D-4F**, blue). Consistent with our binding and *in vitro* neutralization experiments (**Figure 4B-C**), the Group A mAb 3_H2 protected mice infected with the H1N1 vaccine strain but didn’t protect against infection with the drifted H1N1 virus with the K145R substitution (**Figure 4D-4F**, red). Despite receiving the H1/H3 cross- subtypic Group A mAb, mice infected with the drifted H1N1 virus lost weight, displayed clinical signs, and died by 14 days after infection. These data suggest that recent H1N1 viruses have evolved to escape the potent neutralizing activity of the H1/H3 cross-subtype antibodies identified in our study.

## Discussion

Humans are typically infected with influenza viruses early in childhood and continue to encounter antigenically distinct strains throughout life. Prior exposures can impact the specificity of antibody responses to new viral strains. For example, the fine-specificity of antibodies elicited by the 2009 pandemic H1N1 virus in humans was found to be largely dependent on the specific types of seasonal H1N1 viruses that different birth cohorts encountered in childhood^51,52,41,42,53^. Less is known about how prior influenza virus exposures affect antibody responses elicited by entirely different influenza virus subtypes. Here, we identified humans born in the 1990s with high levels of antibodies that cross-react to the variable globular HA head of H3N2 viruses that circulated when these individuals were children and contemporary H1N1 viruses. We found that ferrets sequentially infected with an H3N2 virus from the 1990s and then vaccinated with a contemporary vaccine elicited the same types of antibodies that we identified in humans. Many of these cross-reactive antibodies can potently neutralize virus infections, and H1N1 viruses have started to evolve within the human population to escape this neutralizing activity that targets the HA head near the receptor binding site. Taken together, our data suggest that H3N2 viral exposures in childhood primed long-lived B cell responses that can be later recalled by H1N1 viruses later in life.

The H1/H3 cross-reactive antibodies in our study target an epitope involving residue 145 near the receptor binding site (**Figure 2 and Figure S3**). This is surprising since there is significant amino acid variation in epitopes adjacent to the HA receptor binding site of H1N1 and H3N2 viruses. Residue 145 in H3N2 has alternated between various amino acids over the past 50 years. Notably, the transition between asparagine and lysine at HA residue 145 led to a major antigenic cluster change in H3N2 in the 1990s^48^. Seasonal H1N1 viruses that circulated before 2009 mostly possessed HAs with S145, while the 2009 pandemic H1N1 virus possessed an HA with K145. Given this history, many individuals born in the 1990s were likely first exposed to (i.e., ‘imprinted’ with) an H3N2 virus with an HA with K145 in childhood and later exposed to H1N1 viruses with an HA with K145 after 2009. Although some of the epitope residues adjacent to the receptor binding domains of H1N1 and H3N2 viruses are distinct, it appears that there is sufficient structural homology in the epitope surrounding residue 145, as well as the receptor binding site, that allowed B cells first elicited by 1990s H3N2 exposures to be recalled by exposure to pandemic H1N1 viruses.

Although our data suggest that the cross-reactive H1/H3 B cells identified in our study were primed by H3N2 exposures in the 1990s and boosted by contemporary H1N1 viruses, we cannot exclude the possibility that other types of influenza virus exposures could have contributed to the development of these cross-subtype clones. It is possible that some H1/H3 cross-reactive antibodies can be elicited by primary H1N1 exposures. We found that children with limited exposure histories possessed low levels of H1 antibodies that cross-reacted to an H3 HA from the 1990s; however, individuals born in the 1990s possessed higher levels of these antibodies (**Figure 3A-B**).

The cross-reactive H1/H3 neutralizing antibodies identified in our study likely contributed to protection from pandemic H1N1 infections; however, recent H1N1 viruses have acquired a K145R substitution that prevents binding of these antibodies that are dominant in a subset of individuals born in the 1990s. It is therefore critical for H1N1 vaccine strains to be updated with an HA antigen that possesses R145. The 2023-2024 through the 2025-2026 H1N1 vaccine component was updated with an A/Victoria/4897/2022 HA with R145; however, the high-growth reassortant version of this vaccine strain (A/Victoria/4897/2022 IVR-238) acquired an R145K substitution when it was adapted for growth in eggs. Therefore, the H1N1 component of current egg-based influenza vaccines is mismatched at HA residue 145. Future studies should evaluate whether current egg-propagated and cell-propagated influenza vaccines, which differ at residue 145 of H1 HA, elicit different H1N1 responses in individuals born in the 1990s.

It will also be important to determine if cross-subtype antibodies targeting other epitopes within the HA head domain are prevalent in different birth cohorts. McCarthy et al. characterized B cell responses from 3 human donors born between 1975 and 1979 and identified several H1/H3 mAbs that targeted the globular head of HA^29^. These antibodies targeted an epitope distinct from that recognized by the cross-reactive mAbs in our study, including a footprint involving HA residue 133. Unlike the antibodies identified in our studies, these antibodies were unable to bind to the HA from 2009 pandemic H1N1 viruses, but instead to a subset of older H1N1 viruses that circulated between 1995 and 2006. It is tempting to speculate that the antibodies from the McCarthy study were primed by sequential H3N2 and H1N1 exposures, and this raises the possibility that H1/H3 cross-subtypic binding HA head antibodies may be more common than previously recognized. Future studies should systematically evaluate the presence of these antibodies in distinct birth cohorts and determine if contemporary viruses are evolving to escape these cross-subtypic antibodies. It will also be important to determine if H1/H3 cross-subtypic antibodies targeting the HA head are differentially elicited by initial immune imprints established by infections versus vaccinations^45,54^. Finally, future studies should continue to evaluate the specificity of antibodies elicited in individuals of different ages with distinct influenza virus exposure histories. A greater understanding of how prior influenza virus encounters impact human vaccine responses will ultimately improve vaccine strain selection and vaccine effectiveness.

## Methods

### Human subject recruitment and sampling

All experiments involving humans were approved by the Institutional Review Board of the University of Pennsylvania. Blood samples from 40 adults were collected at the time of vaccination with the 2021-2022 Northern Hemisphere FluLaval Quadrivalent influenza virus vaccine (GlaxoSmithKline) and 7, 28, and 90 days after vaccination between October and December 2021. Plasma and peripheral blood mononuclear cells (PBMCs) were isolated from whole blood for downstream assays. For our experiments, we analyzed sera samples collected 28 days after vaccination and PBMCs collected 7 days after vaccination.

Blood samples from 41 participants were collected as part of longitudinal surveillance of a cohort of households with children, as previously described^55^. Children born in or after 2004 who contributed at least one blood sample between September and December 2021 were eligible for inclusion in this analysis; in those who reported receiving an influenza vaccine, a blood sample collected at least 24 days after vaccination was included.

### Monoclonal antibody production and purification

PBMCs were isolated from whole blood using SepMate™ tubes (Stemcell Technologies) and Histopaque-1077 (Sigma-Aldrich) according to the manufacturer’s instructions. Following PBMC isolation, B cells were enriched using the EasySep™ Human Pan-B Cell Enrichment Kit (Stemcell Technologies) via negative selection. For surface staining, the enriched B cell population was first incubated with a viability dye (Live/Dead^TM^ Fixable Aqua Dead Cell Stain, Invitrogen) in PBS. After washing, cells were stained in FACS buffer (PBS supplemented with 1% BSA and 2 mM EDTA) using the following fluorophore-conjugated antibodies: APC/Cy7- CD19, APC-CD38, PE/Cy7-CD21, PE-CD71, BV421-CD138, BV785-IgD, BV605-CD27, FITC-CD20, and PE/Cy5-conjugated DUMP channel antibodies (CD3, CD14, CD16, CD235a) all from BioLegend or BD Biosciences. Plasmablasts were sorted as live, DUMP⁻, CD19⁺ CD27⁺ CD38⁺ CD20⁻ IgD⁻ cells. Single plasmablasts were sorted into 96-well PCR plates containing catch buffer (1 M Tris-HCl, pH 8.0, with RNase inhibitor (Invitrogen)). Plates were immediately frozen at −80 °C overnight before downstream processing. Complementary DNA (cDNA) was synthesized from single sorted cells using the SuperScript III First-Strand Synthesis System (Invitrogen), following the manufacturer’s protocol. Amplification of immunoglobulin heavy and light chain variable regions was performed using nested multiplex PCR with AmpliTaq Gold DNA Polymerase (Thermo Fisher Scientific) and a validated primer set targeting human immunoglobulin gene families. PCR products were Sanger sequenced. Clonal relationships for antibodies derived from the same donor were assigned based on identical IGHV/GHJ gene usage and similar CDR3 nucleotide sequences (85% similarity); mAbs fulfilling these criteria were annotated as members of the same clone. Heavy and light chain sequences were cloned into IgG1 heavy chain, kappa or lambda light chain constructs (synthesized by Twist). Paired heavy and light chains were transfected into 293F suspension cells (Thermo Fisher) at a density of 1 × 10^6^ cells/mL at a heavy chain to light chain ratio of 1:1 using 293Fectin (Thermo Fisher) in Opti-MEM. Cell supernatants were harvested after four days and clarified by centrifugation.

Antibodies were first concentrated using 30,000 kDa Amicon centrifugal filters (Millipore) and purified with protein G agarose according to the manufacturer’s instructions (Thermo Fisher). IgG was eluted with IgG elution buffer (Thermo Fisher) and neutralized with 1.0 M Tris, pH 8.8. Antibodies were buffer exchanged into DPBS using Amicon centrifugal filters (Millipore).

### Recombinant protein expression and purification for binding assays

Recombinant full-length HA proteins and headless HA proteins were used as antigens in Luminex binding assays and absorption assays. For full-length HA proteins, codon-optimized HA sequences were inserted into pCMV-Sport6 vectors, and HA transmembrane domains were replaced with the trimerization domain from T4 fibritin (foldon), an AviTag, and a hexahistidine tag as previously described^56^. We obtained plasmids encoding recombinant headless H1 and H3 HA proteins from A. McDermott and B. Graham from the Vaccine Research Center at the National Institutes of Health^57,58^. A/New York/631/1996 with the HA K145N mutation was made using the QuikChange II XL site-directed mutagenesis kit (Agilent). HA plasmids were co-transfected with a plasmid encoding neuraminidase (NA) from A/Puerto Rico/8/1934 into 293F suspension cells (Thermo Fisher) at a density of 1 × 10^6^ cells/mL at 1 µg/mL and supernatants were isolated 4 days later and clarified by centrifugation before purification by Ni-NTA affinity chromatography (Qiagen). Purified HAs were buffer exchanged into DPBS using Amicon centrifugal filters (Millipore).

### HA multiplex binding assays

Full-length HA or headless HA proteins were coupled to MagPlex beads (Luminex) at a molar ratio of 0.1 nmol antigen to 1 x 10^6^ beads. 2,500 antigen-coupled beads of each of the antigens were added to each well in 96-well black, clear-bottom plates. Blank beads were included as a negative control. Samples or control monoclonal antibodies were added to the wells before incubating on a microplate shaker at 600 rpm at room temperature for one hour. Plates were washed twice with PBS-TBN (1X PBS, 0.1% bovine serum albumin, 0.02% tween 20, and 0.05% sodium azide), and antigen-specific IgG was detected with mouse anti-human IgG-PE at 2 µg/mL (SouthernBiotech). After 30 minutes incubation, the plates were washed twice with PBS-TBN and read on xMAP INTELLIFLEX (Luminex). All mean fluorescent intensity values were subtracted with blank bead values. Bead mixtures, serum dilution, and secondary antibody dilution were prepared in PBS-TBN.

### Negative-stain EM analysis

Each IgG was incubated with recombinant HA protein (H1 A/California/07/2009 or H3 A/New York/631/1996) at a 1:2 molar ratio (IgG:HA trimer) (n.b. the ratio was experimentally determined to produce well-defined complexes and minimize crosslinking and immune complex formation) for 5 minutes at room temperature prior to grid preparation. Carbon-coated 400-mesh copper grids (Electron Microscopy Sciences) were glow discharged for 25 s at 15 mA using a PELCO easiGlow instrument (Ted Pella, Inc.). IgG–HA complexes were diluted to ∼20 μg/mL in TBS, applied to the grids, and incubated for 30 seconds. After blotting, grids were stained twice with 2% (w/v) uranyl formate for 15 and 60 seconds, respectively. Excess stain was removed by blotting, and the grids were air-dried for at least 5 minutes before imaging. Micrographs were acquired using a 120 kV FEI Tecnai Spirit T12 transmission electron microscope equipped with an FEI Eagle 4k × 4k CCD camera, at a nominal magnification of 52,000×, defocus of –1.5 μm, total electron dose of 25 e⁻/Å², and pixel size of 2.06 Å. Micrographs were collected using Leginon^59^. Particles were picked using a Difference of Gaussians (DoG) particle picker^60^. 2D classification and 3D reconstruction were performed using RELION 3.0^61,62^. Composite maps were generated using UCSF Chimera^61,63^.

### Expression and purification of recombinant HAs and Fabs protein for structure determination

The sequence of influenza A/New York/631/1996 H3N2 HA was obtained from the GISAID database^64^. The ectodomain of A/New York/631/1996 H3N2 HA sequence, which corresponds to 11-329 (HA1) and 1-176 (HA2) based on H3 numbering, was fused with an N-terminal gp67 secretion signal peptide and a C-terminal TEV cleavage site, followed sequentially by a BirA biotinylation site, thrombin cleavage site, trimerization sequence, and a hexahistidine tag. The fusion gene was subsequently cloned into the pFastbac-1 expression vector. The ectodomain construct of influenza A/California/04/2009 H1N1 HA was from the laboratory stock, designed as previously reported^65^. Recombinant bacmid DNA encoding HA ectodomain gene was generated using a Bac-to-Bac baculovirus expression system. Baculovirus was produced by transfecting purified bacmid DNA into Sf9 cells (Thermo Fisher Scientific) using FuGene HD (Promega). Recombinant soluble HA proteins were expressed in High Five cells (Thermo Fisher Scientific) with an MOI of 5-10 for each recombinant virus. After 72 hours at 28°C with shaking at 110 rpm, soluble HA proteins were harvested from supernatant and then further clarified by centrifugation. Soluble HA proteins were purified using Ni-NTA (Qiagen) affinity chromatography, followed by buffer exchange into Tris-buffer saline (TBS; 20 mM Tris, pH 8.0, 300 mM NaCl). The HA ectodomain was digested with trypsin protease (New England Biolabs) during overnight dialysis to remove the C-terminal fusion tags and yield mature cleaved HA proteins. The digested HA proteins were then concentrated and further purified by size-exclusion chromatography using a Hiload 16/90 Superdex 200 column (Cytiva) in TBS.

The head domain constructs (HA1: 43-309, with H3 numbering) of the two subtypes of HAs were subcloned into the pFastbac-1 expression vector with N-terminal gp67 secretion signal peptide and C-terminal hexahistidine tag. Expression procedure of the head domain followed those used for the ectodomain. Head domain proteins in clarified supernatant were purified using Ni Sepharose excel resin (Cytiva) and buffer-exchanged into TBS with 300 mM NaCl. Eluted proteins were concentrated and subjected to size-exclusion chromatography on a Hiload 16/90 Superdex 200 column (Cytiva) in TBS.

The heavy and light chains of 3_H2 and 49_C09 Fabs were cloned into phCMV3 mammalian cell expression vector, with an N-terminal IgK secretion peptide. The heavy and light chain constructs were co-transfected into the ExpiCHO cells (Thermo Fisher Scientific) according to the manufacturer’s protocol. Fabs were purified using the CH1-XL affinity matrix (Thermo Fisher Scientific) to capture the CH1 constant domain and followed by buffer exchange into PBS (pH 7.4). Final purification was performed by size-exclusion chromatography in TBS (pH 8.0).

### Crystallization and structural determination

Fab 3_H2 was incubated with purified A/California/04/2009 HA trimer and head domain at a molar ratio of 1.2:1 (Fab:HA monomer/HA head) overnight at 4°C, respectively, and the final concentration of HA trimer was about 8 mg/mL and the HA head domain was ∼10 mg/mL. 49_C09 Fab-HA complexes were prepared following the same protocol. Crystallization screening was carried out using our high-throughput, robotic CrystalMation system (Rigaku) under 18°C at TSRI^66^. Initial screening employed the sitting drop vapor diffusion method, with 35 µL reservoir solution and each drop consisting of 0.1 µL complex sample mixed with 0.1 µL precipitant. Diffraction-quality crystals were obtained under the conditions below:

| Complexes | Crystallization conditions | Cryo-protectant |
| --- | --- | --- |
| 3_H2+A/California/04/2009<br>HA | 0.2M Tri-potassium citrate,<br>20% (w/v) PEG3350, pH 8.3 | 10% (v/v)<br>ethylene glycol |
| 3_H2+A/New York/631/1996<br>HA head | 0.2M di-potassium hydrogen phosphate,<br>20% (w/v) PEG3350, pH 9.2 | 10% (v/v)<br>ethylene glycol |
| 49_C09+A/California/04/2009<br>HA | 2M ammonium sulfate, 15% (v/v)<br>ethylene glycol, 0.2M lithium sulfate,<br>0.1M CAPS, pH 10.5 | None |
| 49_C09+A/New York/631/1996<br>HA head | 4M sodium formate | 20% (v/v)<br>Glycerol |
| 49_C09+A/Victoria/2570/2019<br>HA head | 0.1M Tris, pH 8.0, 65% (v/v) MPD | None |

Diffraction data were collected at beamline 12-1 of the Stanford Synchrotron Radiation Lightsource (SSRL), and beamline 17-ID-2 (FMX) of the National Synchrotron Light Source II (NSLS-II) at Brookhaven National Laboratory. Data indexing, integration and scaling were carried out in HKL2000 (HKL Research)^67^. Structures were solved by molecular replacement using Phaser^68^ within the PHENIX software suite^69^, followed by automated refinement with phenix.refine. Manual refinement and modelling were conducted in the Coot^70^. For molecular replacement, PDB 4M4Y and 4HMG were used as models of H1 and H3 HAs, respectively, and the initial model of Fabs were generated by AlphaFold 2^71^. The Kabat numbering scheme was used for the Fabs, and the H3 numbering scheme for HAs. All structural graphs were prepared using PyMOL and UCSF ChimeraX^63^.

### Carboxyl magnetic bead absorption assays

Full-length HA proteins were coupled to carboxyl magnetic beads (Raybiotech) at a concentration of 35 µg antigen per 100 µL magnetic beads. As negative controls, mock beads were prepared by adding DPBS in place of antigens. Antigens or DPBS were added to magnetic beads and incubated at 4°C with rotation for two hours. The unbound fractions were removed using a magnetic stand, and beads were quenched by incubating in 50 mM Tris, pH 7.4 at room temperature with rotation for 15 minutes. Quenching buffer was removed using the magnetic stand, and the coupled beads were washed 4 times with wash buffer (DPBS with 0.1% BSA and 0.05% tween 20). Beads were resuspended in 1 bead-volume of wash buffer before storage at 4°C.

For absorptions, sera were first diluted 1:80 with sterile DPBS and then brought to 1:100 dilution with the addition of the coupled beads at a final bead:diluted sera volume ratio of 1:4.

The bead/sera mixture was vortexed and placed on a shaker at 2000 RPM for one hour at room temperature. Bead-sera mixtures were then placed on a magnet for separation and unabsorbed fractions were removed and transferred to a clean tube.

### Viruses

All influenza viruses in this study were generated by reverse-genetics (MJB 45). HA and NA genes from A/Victoria/2570/2019, A/New York/631/1996, and A/Minnesota/35/2022 were cloned into the pHW2000 reverse-genetics plasmid. A/Minnesota/35/2022 with a Q226R mutation in the HA gene was generated using the QuikChange II XL site-directed mutagenesis kit (Agilent). All viruses were launched with their respective HA and NA genes and internal segments from A/Puerto Rico/8/1934 H1N1. Each of the eight segments was co-transfected with Lipofectamine 2000 (Thermo Fisher) in a co-culture of 293T and Madin-Darby Canine Kidney (MDCK)-SIAT1 cells. Transfection supernatants were harvested three days after transfection and used for virus expansion. A/Victoria/2570/2019 and A/Minnesota/35/2022_Q226R were further propagated in embryonated chicken eggs. A/New York/631/1996 and A/Minnesota/35/2022 were expanded in MDCK-SIAT1 cells.

### Focus reduction neutralization test (FRNT)

96-well flat bottom tissue culture plates were seeded with 2.5 x 10^4^ MDCK-SIAT1 cells per well in Minimum Essential Media (MEM; Gibco) supplemented with 10% FBS the day before. Serum samples or mAbs were serially diluted in serum-free MEM and mixed with a virus dilution that resulted in ∼300 focus forming units (FFU) per well in virus-only control wells and incubated at room temperature for one hour. Cells were washed twice with serum-free MEM and virus/antibody mixtures were transferred to the 96-well flat bottom plate and incubated at 37°C in 5% CO_2_ for one hour. After incubation, cells were washed once with serum-free MEM and an overlay of 1.25% Avicel (IFF Pharma Solutions) in MEM supplemented with 0.2% gentamicin (Gibco) and 1% 1 M HEPES (Gibco) was added to the cells. The cells were incubated for 18 hours at 37°C in 5% CO_2_, after which the overlay was removed and the cells were fixed with 4% paraformaldehyde for one hour at 4°C in the dark. PFA was removed, and 0.5% Triton X-100 in DPBS were added to permeabilize the cells for seven minutes. Triton was removed from wells, followed by blocking with 5% milk in DPBS for one hour at room temperature. Plates were washed five times with distilled water and mouse anti-nucleoprotein antibody (clone IC5–1B7) diluted in 5% milk in DPBS was added to each well. After incubation for one hour at room temperature, plates were washed five times and goat anti-mouse peroxidase-conjugated secondary antibody (SouthernBiotech) diluted in 5% milk in DPBS was added to each well.

Plates were incubated for one hour at room temperature, washed five times, and then TrueBlue peroxidase substrate (SeraCare) was added to the plates for one hour in the dark. Substrate was removed and plates were scanned on the ImmunoSpot S6 Analyzer with the bioSpot program (CTL) for visualization and enumeration of the foci. IC_50_ are reported as the highest reciprocal serum dilution or the lowest antibody concentration that inhibited at least 50% of virus in relation to virus-only control wells. Serum samples or mAbs that reduced the amount of virus less than 50% at 20 µg/mL were assigned an IC_50_ of 40 µg/mL.

### Ferret experiments

Ferret experiments were completed at the University of Pittsburgh in compliance with the guidelines of the Institutional Animal Care and Use Committee under approved protocol 22061230. Six-month-old, immunologically naïve, male ferrets were purchased from Triple F Farms (Sayre, PA). Ferrets were intranasally inoculated with 100,000 TCID_50_ A/Nanchang/933/1995 or 100,000 TCID_50_ A/Nanchang/933/1995_N145K in a total volume of 500 µL split between nostrils. Ferrets were vaccinated intramuscularly (IM) 84 days after challenge with 250 µL 2022-2023 FluLaval quadrivalent influenza vaccine. Control group only received the vaccine. Ferret sera were collected throughout the study. The sample size was determined based on similar previous experiments.

### Mouse experiments

Female DBA/2J mice (Jackson Laboratories) aged 7 to 8 weeks were injected with 100 µg of mAbs in 100 µL DPBS intraperitoneally (IP). As controls, some mice received an intraperitoneal injection of 100 µL DPBS before infection. Mice were anesthetized by inhalation of isoflurane (4 to 5% in oxygen) and intranasally inoculated with 8,000 TCID50 A/Victoria/2570/2019 or 8,000 TCID50 A/Minnesota/35/2022 in 50 µL of DPBS six hours after IP injection.

A/Minnesota/35/2022 virus possessed a Q226R HA mutation that increases infectivity in mice. We monitored mice daily for lethargy, hunched posture, ruffled fur, and labored breathing. Clinical severity was scored by an analyst blinded to the experimental groups. We also monitored weight loss and survival. All murine experiments were approved by the Institutional Animal Care and Use Committees of the Wistar Institute and the University of Pennsylvania.

### Phylogenetic reconstruction of H3N2 and pandemic H1N1 sequences

To contextualize the HA sequences of H3N2 strains between 1968 and 2006 and H1N1 strains between 2009 and 2025, phylogenetic trees were constructed using the Nextstrain pipeline^72^. All available full-length HA sequences from human H3N2 isolates collected between 1975 and Dec 31, 2006, and human H1N1 isolates collected between October 1, 2009, and April 30, 2025, were downloaded from GISAID (gisaid.org). The sequences were randomly subsampled to 50 isolates per month to build the H3N2 tree and 20 isolates per month to build the H1N1 tree. Sequences were aligned with MAFFT^73^ and divergence phylogenies were created using IQ- TREE 2^74^. Tree visualization was performed using Baltic (github.com/evogytis/baltic). Frequencies of amino acids at residue 145 were estimated with Augur^75^ using the KDE estimation method. Frequency was visualized with Auspice^72^.

### Quantification and statistical analysis

All statistical tests were performed in R version 4.3.1, and data were visualized using custom scripts in R Studio. Sample sizes were determined based on sample availability. Wilcoxon rank-sum or signed rank tests were performed depending on unmatched or matched samples. Benjamini-Hochberg corrections were made for multiple comparisons. Statistical significance was defined as P < 0.05.

## Data availability

The X-ray coordinates and structure factors are deposited in the Protein Data Bank (PDB) with PDB codes 9Y3X for 3_H2-A/California/04/2009 HA, 9Y3Z for 3_H2-A/New York/631/1996 HA, 9Y3Y for 49_C09-A/California/04/2009 HA, 9Y40 for 49_C09-A/New York/631/1996 HA, and 9Y41 for 49_C09-A/Victoria/2570/2019 HA. Negative-stain EM maps of A/California/07/2009 H1N1 HA in complex with antibodies 97_F7, 88_B4, 3_H2, 49_C09, 33_C08, 33_C02, and 18_D11, as well as A/New York/631/1996 H3N2 HA in complex with antibodies 97_F7, 88_B4, 33_C08, and 18_D11, have been deposited in the Electron Microscopy Data Bank (EMDB) under accession codes EMD-72527 to EMD-72537.

## Acknowledgement

We would like to thank the study participants for their generosity in making the study possible. We gratefully acknowledge labs that have generated sequence data deposited into public databases. We also thank Penn’s Immune Health team for processing human samples and members of the Hensley lab for helpful discussions and feedback. We thank Henry Tien for helping with the automated robotic crystal screening at TSRI, and Wenli Yu for assistance in the protein expression and purification. We are also grateful to the staff of SSRL and NSLS-II for their assistance in X-ray data collection. Molecular graphics and analyses performed with UCSF ChimeraX, developed by the Resource for Biocomputing, Visualization, and Informatics at the University of California, San Francisco, with support from National Institutes of Health R01-GM129325 and the Office of Cyber Infrastructure and Computational Biology, National Institute of Allergy and Infectious Diseases. This project has been funded with Federal funds from the National Institute of Allergy and Infectious Diseases, National Institutes of Health, Department of Health and Human Services, under Contract No. 75N93021C00015 (S.E.H., S.D.B., I.A.W., A.B.W., S.S.L., E.T.M.).

## Author contributions

Conceptualization: S.H.L., S.D.B., and S.E.H.

Methodology: S.H.L., B.W., G.J., and V.L.

Investigation: S.H.L, B.W., G.J., A.M., R.K.A., V.L., C.F., J.T.O., N.Y., K.R., S.A.J., J.L., G.H.J., T.A.P.

Human sample collection: E.M.D., A.P.C., R.G.C., A.S.M., E.T.M.

Formal analysis: S.H.L., B.W., G.J.

Visualization: S.H.L., B.W., and G.J.

Writing—original draft: S.H.L. and S.E.H.

Writing—review and editing: all authors

Supervision: J.H., S.S.L., A.B.W., I.A.W., S.D.B., S.E.H.

Funding acquisition: S.E.H., S.D.B., I.A.W., A.B.W., S.S.L., A.S.M., E.T.M.

## Declaration of interests

S.E.H. is a co-inventor on patents that describe the use of nucleoside-modified mRNA as a vaccine platform. S.E.H reports receiving consulting fees from Sanofi, Pfizer, Lumen, Novavax, and Merck. S.D.B. has consulted for Regeneron, Sanofi, Novartis, Genentech, Visterra, and Janssen on topics unrelated to this study, is a scientific co-founder of Immunera Inc., and owns stock in AbCellera Biologics Inc.

---

**Figure S1. Binding of 2021-2022 inactivated influenza vaccine-elicited antibodies to a historical panel of influenza HAs.** (**A**) Monoclonal antibodies isolated seven days after influenza vaccination were measured against the HAs of the H1N1, H3N2, and IBV vaccine components (A/Victoria/2570/2019, A/Tasmania/503/2020, B/Washington/02/2019, and B/Phuket/3073/2013 respectively; bolded), a panel of historical H1, H3, and IBV HAs, and group 1 and group 2 headless HA constructs in a multiplexed binding assay. The intensity of binding is shown in MFI. (**B**) Proportion of 2021-2022 influenza vaccine-elicited antibodies that react to each of the vaccine components is shown. Data represent the mean of two independent experiments.

**Figure S2. H3 amino acid identity at HA residue 145 between 1968 and 2006.** H3N2 phylogenetic tree shows amino acid variation at HA position 145. Dark blue tips indicate that viruses had lysine (K) at residue 145, and grey tips indicate viruses with amino acids other than lysine at residue 145.

**Figure S3. Epitope mapping and comparison of Group A mAb 3_H2 and Group B mAb 49_C09.**

**(A)** Crystal structure of the A/Victoria/2570/2019 H1 HA-Fab 49_C09 complex. The HA head domain is shown in surface representation in grey color, with the epitope regions (defined as residues with a BSA > 0 Å^2^) highlighted in turquoise. Antibodies are shown in cartoon representation, with heavy chains colored in dark red and light chains colored in wheat. Labeled residues indicate those involved in hydrogen bonds or salt bridge formation upon HA engagement. Epitope mapping and comparison of 3_H2 (**B**) and 49_C09 (**C**) mAbs on H1 and H3 HAs. In the upper sequence alignments, epitope residues are highlighted, and the darker color indicates residues involved in hydrogen bonds or salt bridges with the antibodies. The corresponding epitope surfaces are displayed on the structures and outlined with dark lines. (**D**) BSA contributions (Å^2^) of the heavy and light chains in the various HA complex structures.

**Figure S4. Polyclonal H1/H3 cross-reactive antibody responses after vaccination in humans and ferrets.** (**A**) Serum binding to H1 vaccine HA (A/Victoria/2570/2019) was measured after incubating with HA-coupled beads to absorb antibodies reactive to 1996 H3 HA with HA K145 (A/New York/631/1996) or 1996 H3 HA with HA N145 (A/New York/631/1996_K145N) in the four individuals with the highest percentage of H1/H3 cross- reactive antibodies in **Figures 3A** and **3B**. Antibody titers are represented as percentages of cross-reactive antibodies. (**B**) Ferrets (n = 4 per group) were infected with A/Nanchang/933/1995 H3N2 with HA K145 (blue) or A/Nanchang/933/1995 with HA N145 (red) and vaccinated 84 days later with a half-dose human FluLaval influenza vaccine. Naïve ferrets only received the vaccine on day 84 (grey). Binding against 1996 H3 with HA K145 (A/New York/631/2009) and 1996 H3 with HA N145 (A/New York/631/2009_K145N) was quantified in a multiplexed binding assay. Data are representative of two independent experiments. Horizontal bars in (**A**) indicate means.

**Figure S5. Amino acid substitutions in H1N1 circulating strain A/Minnesota/35/2022 relative to H1N1 vaccine strain A/Victoria/2570/2019.** Structure of A/California/04/2009 H1 HA (PDB: 4M4Y) with residue K145 highlighted in red on the head domain and additional HA substitutions in A/Minnesota/35/2022 relative to H1 vaccine strain A/Victoria/2570/2019 highlighted in blue.

## Notes

### Author Declarations

All experiments involving humans were approved by the Institutional Review Board of the University of Pennsylvania.

