## Supplemental Figures for "Childhood immunological imprinting of cross-subtype antibodies targeting the hemagglutinin head domain of influenza viruses"

### Figure S1

**A**

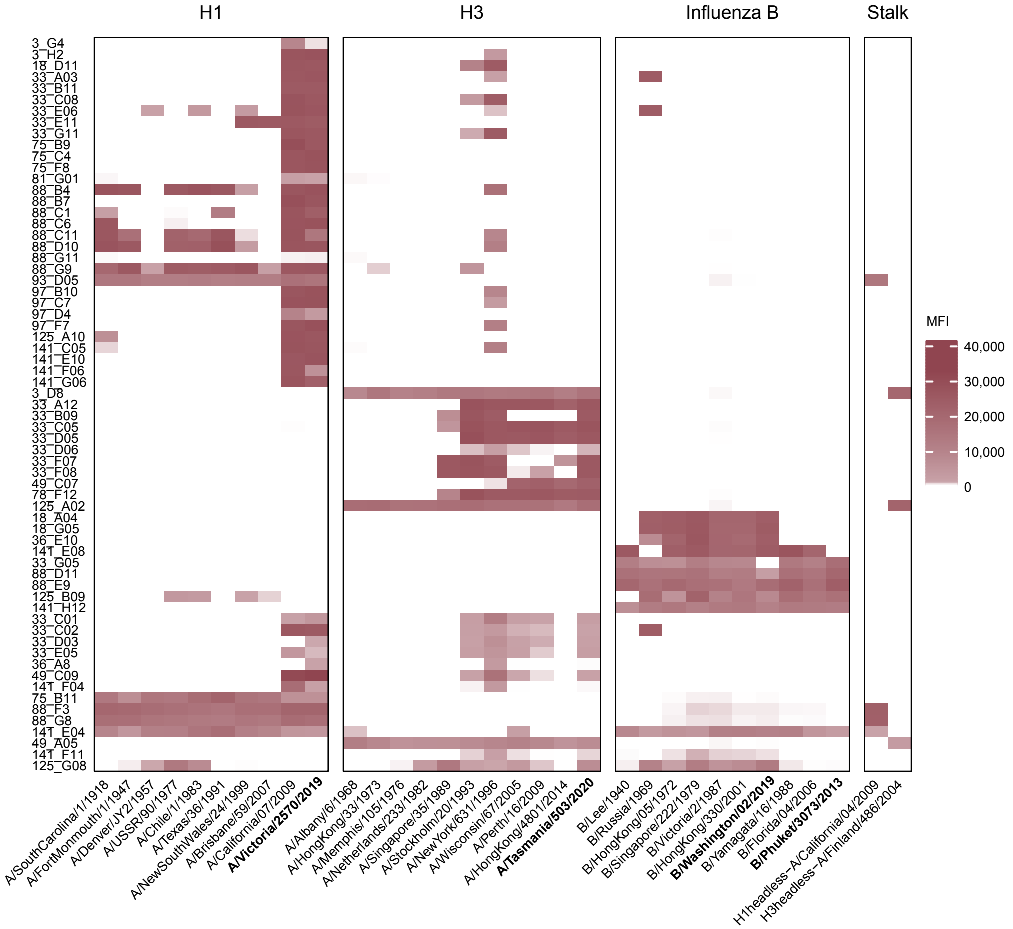
H1
H3
Influenza B
Stalk
3_G4
3_H2
18_D11
33_A03
33_B11
33_C08
33_E06
33_E11
33_G11
75_B9
75_C4
75_F8
81_G01
88_B4
88_B7
88_C1
88_C6
88_C11
88_D10
88_G11
88_G9
93_D05
97_B10
97_C7
97_D4
97_F7
125_A10
141_C05
141_E10
141_F06
141_G06
3_D8
33_A12
33_B09
33_C05
33_D05
33_D06
33_F07
33_F08
49_C07
78_F12
125_A02
18_A04
18_G05
36_E10
141_E08
33_G05
88_D11
88_E9
125_B09
141_H12
33_C01
33_C02
33_D03
33_E05
36_A8
49_C09
141_F04
75_B11
88_F3
88_G8
141_E04
49_A05
141_F11
125_G08
MFI
40,000
30,000
20,000
10,000
0
A/SouthCarolina/1/1918
A/FortMonmouth/1/1947
A/Denver/JY2/1957
A/USSR/90/1977
A/Chile/1/1983
A/Texas/36/1991
A/NewSouthWales/24/1999
A/Brisbane/59/2007
A/California/07/2009
A/Victoria/2570/2019
A/Albany/6/1968
A/HongKong/33/1973
A/Memphis/105/1976
A/Netherlands/233/1982
A/Singapore/35/1989
A/Stockholm/20/1993
A/NewYork/631/1996
A/Wisconsin/67/2005
A/Perth/16/2009
A/HongKong/4801/2014
A/Tasmania/503/2020
B/Lee/1940
B/Russia/1969
B/HongKong/05/1972
B/Singapore/222/1979
B/Victoria/2/1987
B/HongKong/330/2001
B/Washington/02/2019
B/Yamagata/16/1988
B/Florida/04/2006
B/Phuket/3073/2013
H1headless-A/California/04/2009
H3headless-A/Finland/486/2004

**B**

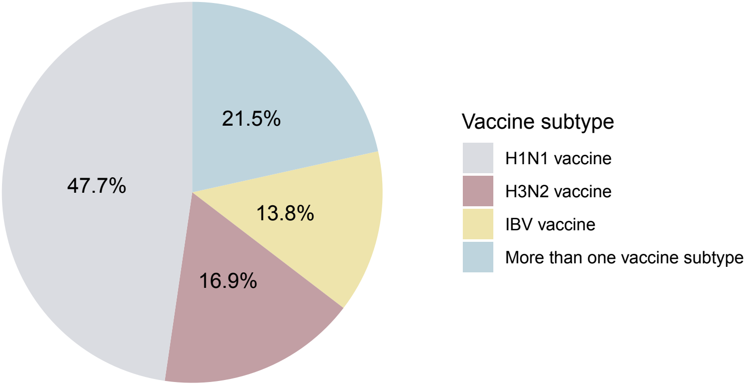
21.5%
47.7%
13.8%
16.9%
Vaccine subtype
H1N1 vaccine
H3N2 vaccine
IBV vaccine
More than one vaccine subtype

### Figure S2

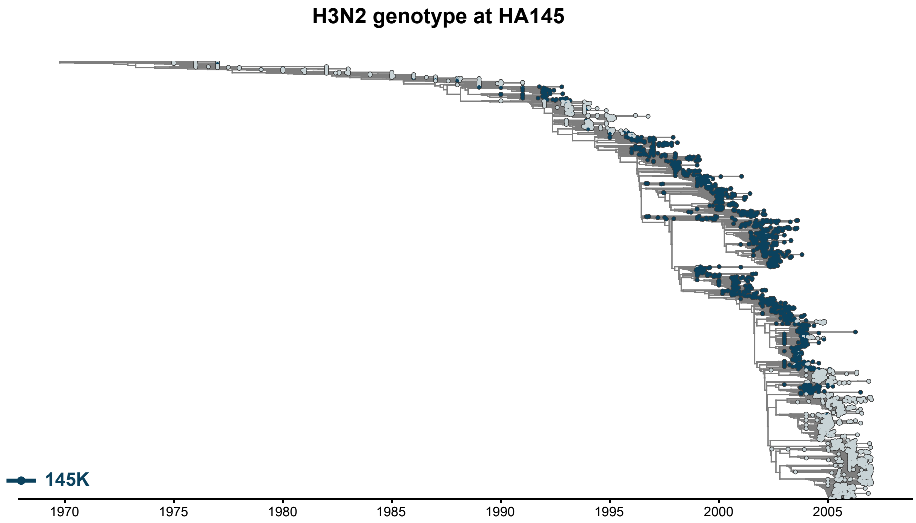

H3N2 genotype at HA145
145K
1970
1975
1980
1985
1990
1995
2000
2005

### Figure S3

**A**

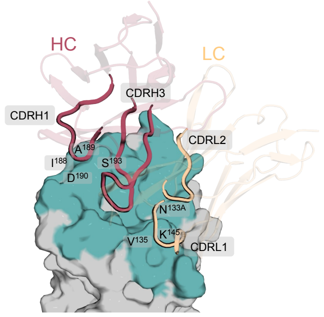


**2019 H1 HA Head + Fab 49_C09**

**B**

**3_H2 epitopes mapping and comparison**

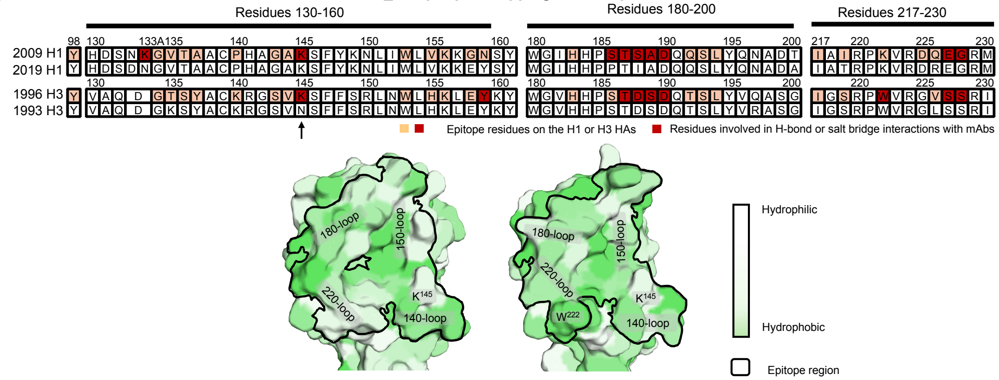


**C**

**49_C09 epitopes mapping and comparison**

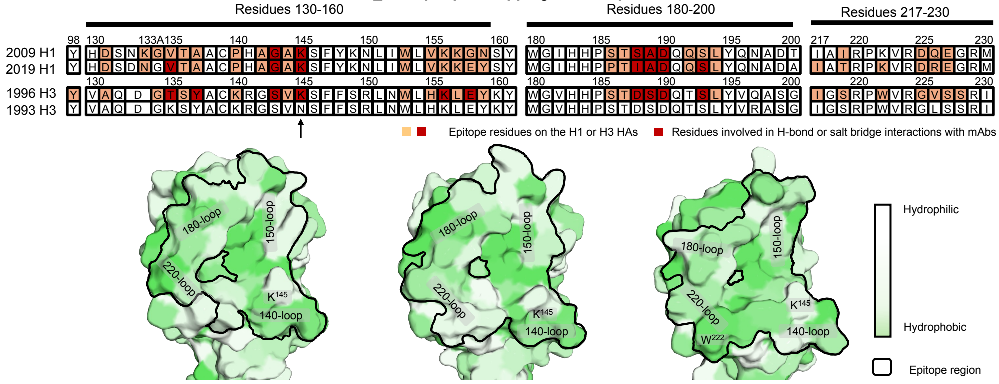


**D**

Buried surface area (BSA, Å²) of 3_H2 in complex structures

| | HC | | | LC | | | |
|---|---|---|---|---|---|---|---|
| | CDRH1 | CDRH2 | CDRH3 | CDRL1 | CDRL2 | CDRL3 | FR3 |
| 2009 H1 HA+3_H2 | 99 | 20 | 404 | 46 | 196 | 38 | 97 |
| 1996 H3 HA+3_H2 | 102 | 9 | 380 | 66 | 126 | 51 | 89 |

Buried surface area (BSA, Å²) of 49_C09 in complex structures

| | HC | | | LC | | | |
|---|---|---|---|---|---|---|---|
| | CDRH1 | CDRH2 | CDRH3 | CDRL1 | CDRL2 | CDRL3 | FR3 |
| 2009 H1 HA+49_C09 | 80 | 15 | 286 | 143 | 293 | 5 | 44 |
| 2019 H1 HA+49_C09 | 101 | 14 | 371 | 148 | 277 | 39 | 46 |
| 1996 H3 HA+49_C09 | 123 | 5 | 437 | 170 | 251 | 0 | 53 |

#### Figure S4

A

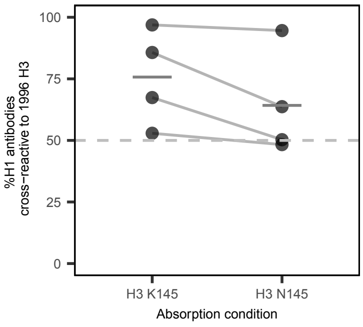

%H1 antibodies
cross−reactive to 1996 H3
100
75
50
25
0
H3 K145
H3 N145
Absorption condition


B

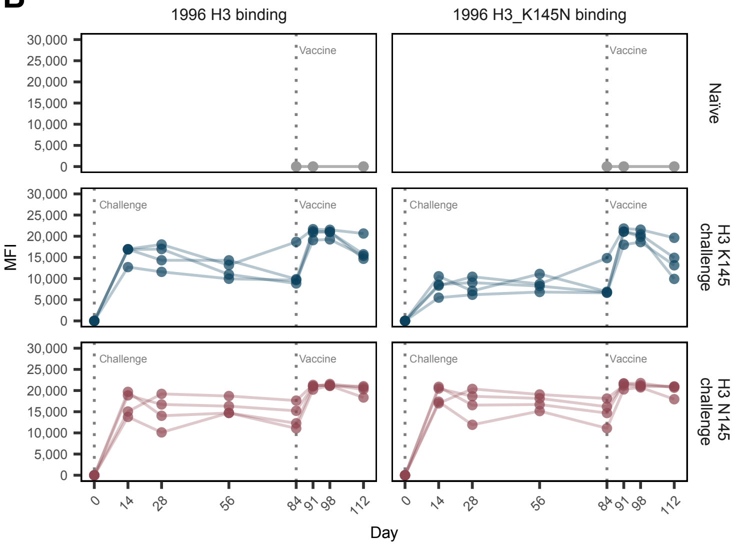

1996 H3 binding
1996 H3_K145N binding
Naïve
H3 K145 challenge
H3 N145 challenge
Challenge
Vaccine
MFI
30,000
25,000
20,000
15,000
10,000
5,000
0
0
14
28
56
84
91
98
112
Day

**Figure S5**

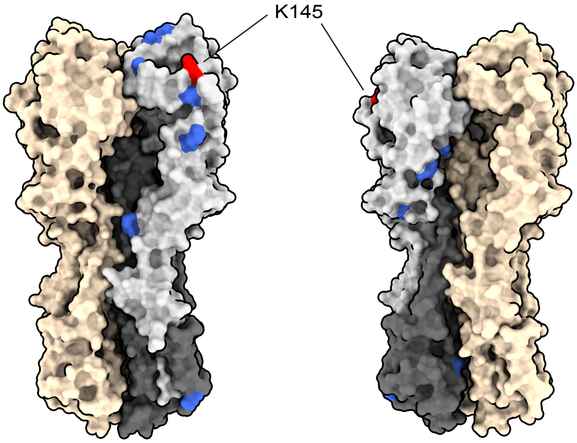
